# Increased GDF11 expression associated with increased survival of Grade 3 Gliomas

**DOI:** 10.1101/2022.08.16.22278840

**Authors:** Steven Lehrer, Peter H. Rheinstein

**Affiliations:** Department of Radiation Oncology, Icahn School of Medicine at Mount Sinai, New York; Severn Health Solutions, Severna Park, Maryland

**Keywords:** glioma, therapy, temozolomide

## Abstract

**Background:** Infusing young mouse blood into old mice makes the old mice biologically younger. When an old mouse and a young mouse share a circulatory system, the old mouse’s muscle function is improved, and the production of olfactory neurons is increased. GDF11 seems to be a crucial element of the young blood in both instances.

**Methods:** Because of GDF11’s potential neuroprotective actions, we used The Cancer Genome Atlas (TCGA) to assess the effect of GDF11 expression in malignant gliomas. We analyzed the GDC TCGA lower grade glioma data set. To access TCGA data we used the Xena platform and cBioportal. Statistical analysis was done with SPSS v26.

**Results:** increased GDF11 expression in IDH1 mutant subjects was significant. There was significantly increased survival (p = 0.00065, log rank test) with high GDF11 expression in grade 3 gliomas. The survival effect was less prominent in grade 2 gliomas. GDF11 gene expression was highest in anaplastic oligodendrogliomas and mixed gliomas with 1p 19q co-deletions and few or no TP53 or ATRX mutations. GDF11 gene expression was lowest in anaplastic astrocytomas with no 1p 19q co-deletions and many TP53 and ATRX mutations.

**Conclusion:** GDF11 or an analogue might be therapeutic in grade 3 glioma. GDF11 does not cross the blood brain barrier but affects the brain by acting on brain endothelial cells. GDF11 might be delivered to a brain tumor intranasally.

---

Growth differentiation factor 11 (GDF11) is a protein that is encoded by the growth differentiation factor 11 gene. GDF11 is a member of the Transforming growth factor beta family.

GDF11 inhibits age-related reductions in both neurogenesis and muscle function. Infusing young mouse blood into old mice makes the old mice biologically younger [1]. When an old mouse and a young mouse share a circulatory system, Sinha et al. discovered that the old mouse’s muscle function is improved [2], while Katsimpardi et al. observed that the production of olfactory neurons is increased [3]. GDF11 seemed to be a crucial element of the young blood in both instances. In older adults, low muscle mass is associated with diminished cognitive function [4].

In the brain, in contrast to its function during development, recombinant GDF11 (rGDF11) increases neurogenesis when given systemically to old mice [5]. It has antiaging effects on skin [6] and other tissues and may be of value in treatment of stroke.

Because GDF11 is highly expressed in the brain and potentially neuroprotective [7], we used The Cancer Genome Atlas (TCGA) to assess the effect of GDF11 expression in malignant gliomas.

## Methods

Over 20,000 primary cancer and matched normal samples from 33 different cancer types are molecularly described by The Cancer Genome Atlas (TCGA). The National Human Genome Research Institute and National Cancer Institute started working together on this project in 2006. Over 2.5 petabytes of genomic, epigenomic, transcriptomic, and proteomic data have been produced by TCGA [8].

To access TCGA data we used the Xena platform [9] and cBioportal [10]. Statistical analysis was done with SPSS v26.

## Results

We analyzed the GDC TCGA lower grade glioma data set. 264 subjects had grade 2 (g2) tumors. 266 subjects had grade 3 (g3) tumors. The mean age of g2 was 40 ± 13, of g3 was 45 ± 13; the age difference was significant (p < 0.001). 45% of the patients were female, 55% were male.

34 subjects had no IDH1 mutation and GDF11 17.02 ± 0.51 log2(fpkm-uq+1), 91 subjects had IDH1 mutation and GDF11 17.3 ± 0.52 log2(fpkm-uq+1). The increased GDF11 expression in IDH1 mutant subjects was significant (p = 0.004, two tailed t test).

Survival probability by GDF11 expression is shown in Figure 1. Note the significantly increased survival (p = 0.00065, log rank test) with high GDF11 expression in grade 3 gliomas. The survival effect was less prominent in grade 2 gliomas.

**Figure 1.**
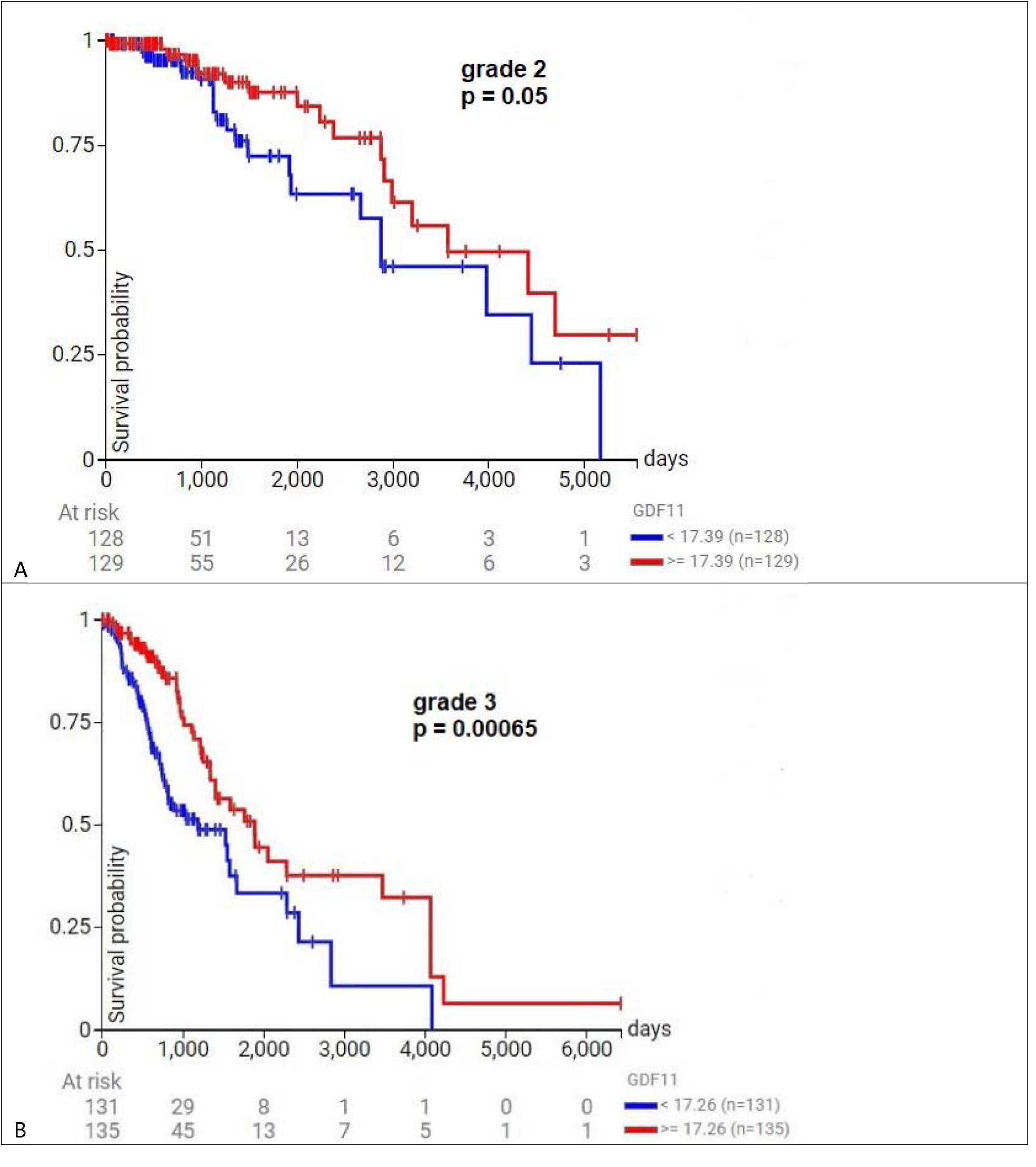
Survival probability in gliomas by GDF11 expression. A) grade 2 glioma, B) grade 3 glioma. Note the significant relationship (p = 0.00065, log rank test) of high GDF11 expression (red curve) with increased survival of grade 3 gliomas. The effect was less prominent in grade 2 gliomas.

Genetic separation of grade 3 gliomas into two disease groups in 257 patients is shown in Figure 2. GDF11 gene expression is highest in anaplastic oligodendrogliomas and mixed gliomas with 1p 19q co-deletions and few or no TP53 or ATRX mutations (81 patients, GDF11 17.58 ± 0.58 log2(fpkm-uq+1)).

**Figure 2.**
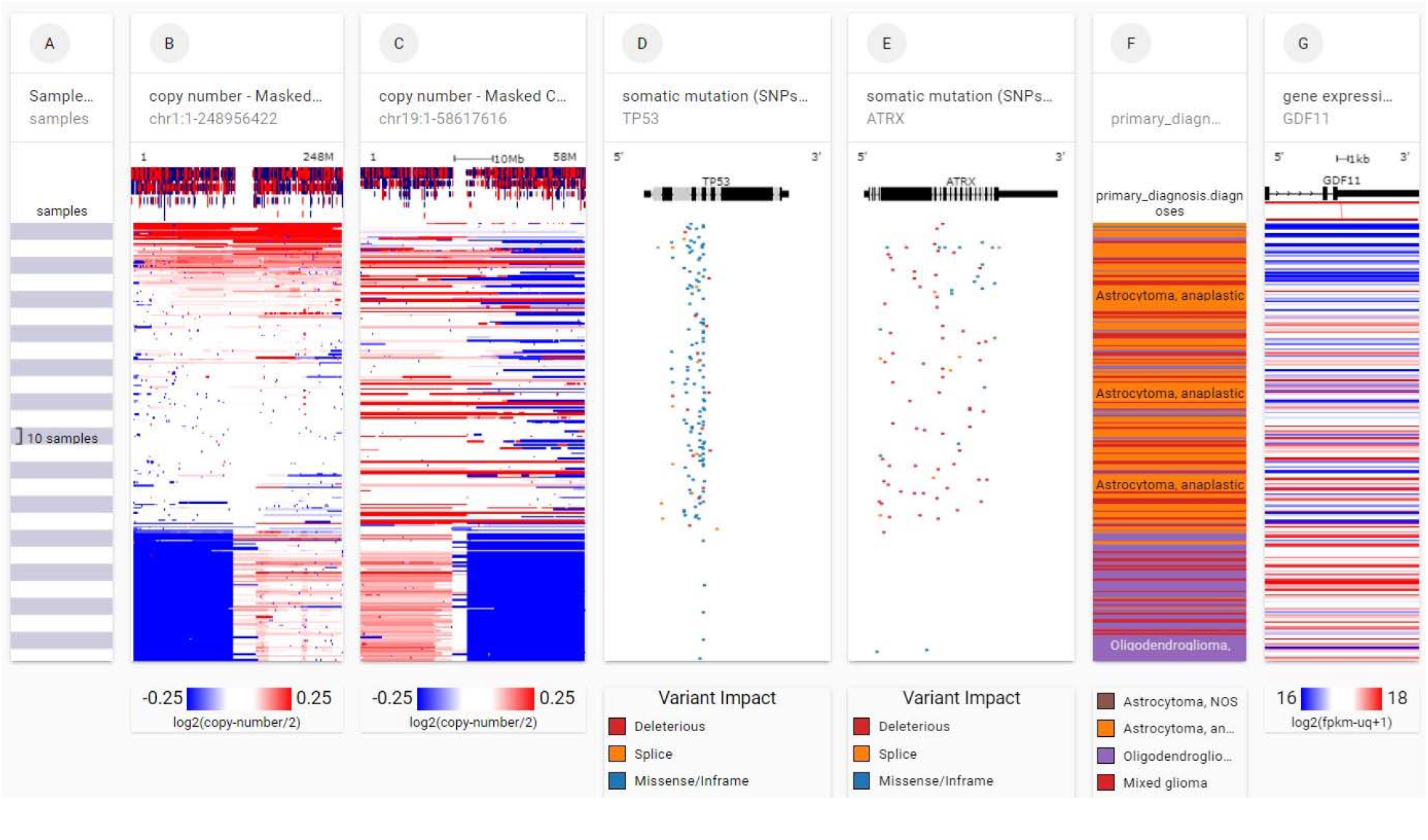
Genetic separation of grade 3 gliomas into two disease groups in 166 patients. Each row contains data from a single sample. Row order is determined by sorting the rows by their column values. Each gray or white band in column A indicates 10 samples. Loss of chromosome arms 1p and 19q are indicated by blue blocks, columns B and C. TP53 and ATRX mutations are indicated in columns D and E. Note that GDF11 gene expression is highest (red bands column G) in anaplastic oligodendrogliomas and mixed gliomas (column F) with 1p 19q co-deletions and few or no TP53 or ATRX mutations, disease group 1. GDF11 gene expression is lowest (blue bands column G) in anaplastic astrocytomas (orange bands column F) with no 1p 19q co-deletions and many TP53 and ATRX mutations, disease group 2 (UCSC Xena http://xena.ucsc.edu).

GDF11 gene expression is lowest in anaplastic astrocytomas with no 1p 19q co-deletions and many TP53 and ATRX mutations (176 patients, GDF11 17.1 ± 0.62 log2(fpkm-uq+1)). The difference in GDF11 expression in the two groups is significant (p < 0.001, two tailed t test).

Only one mutation was present in 257 grade 3 gliomas, a silent mutation. But GDF11 expression was highly correlated with Homeodomain-interacting protein kinase 2 (HIPK2) expression (Figure 3).

**Figure 3.**
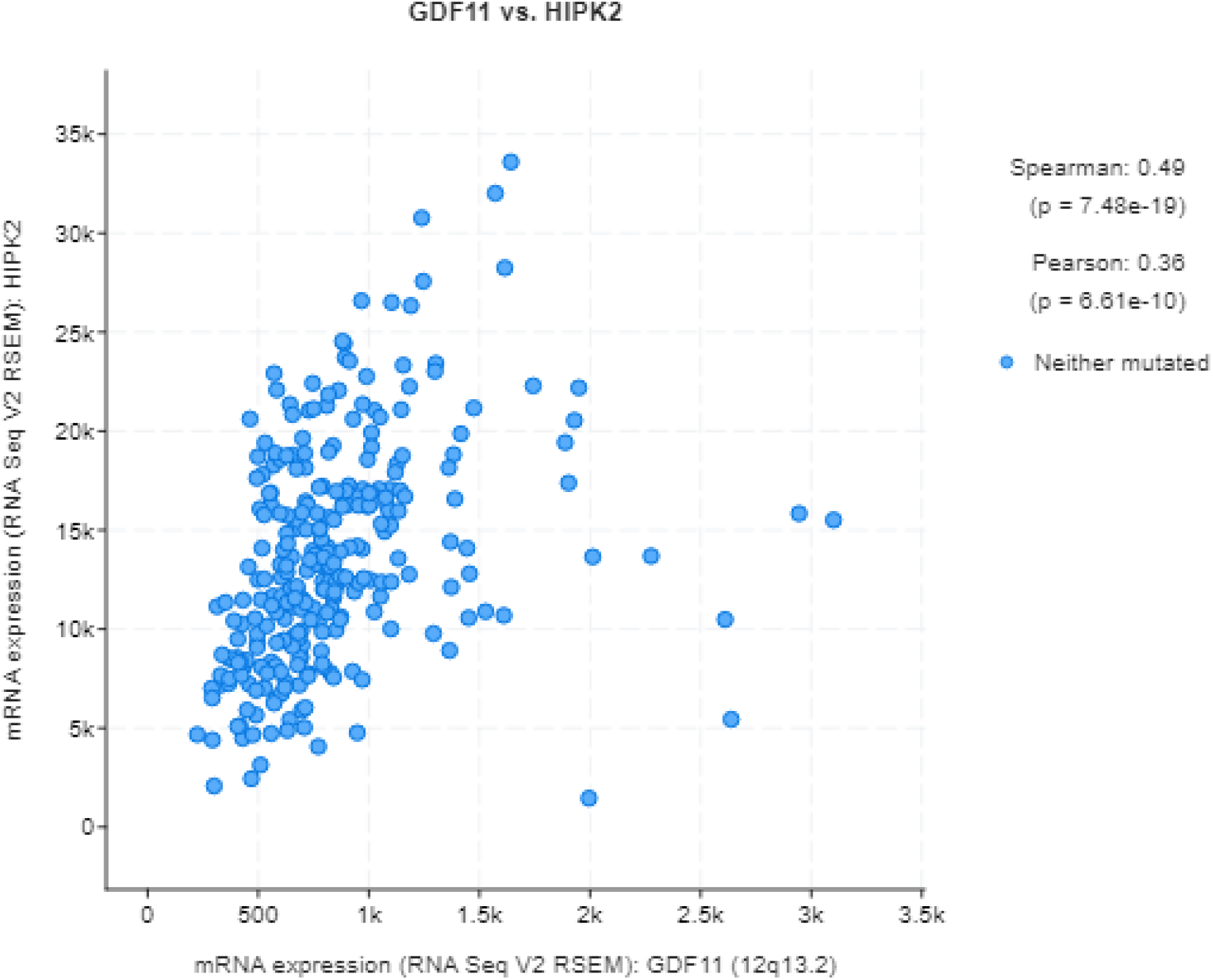
GDF1 expression was highly correlated with HIPK2 expression.

## Discussion

Grade 3 gliomas are divided into two categories according to the 2021 WHO classification: grade 3 IDH mutant (IDHmt), 1p/19q codeleted oligodendroglioma, and IDH mutant astrocytoma. Surgery is performed first, followed by radiation treatment (RT) and chemotherapy with alkylating agents.

Patients with non-codeleted grade 3 IDHmt astrocytomas have better outcomes when treated with RT and temozolomide. Large randomized controlled trials support the use of procarbazine, CCNU, and vincristine in the treatment of patients with IDHmt, codeleted oligodendroglioma. With these measures, patients with grade 3 IDHmt astrocytomas should expect to live an additional 10 years, while those with grade 3 IDHmt codeleted oligodendrogliomas can expect an additional 14 years [11].

IDHmt grade 3 gliomas can be successfully treated with RT and alkylating drugs, but, typically, first therapy is ineffective; IDHmt tumors virtually always return and worsen. Various new treatments are currently under study: IDH inhibitors, immunotherapy, and poly (ADPRibose) polymerase (PARP) inhibitors that disrupt DNA damage repair pathways.

Homeodomain-interacting protein kinase 2 (HIPK2) regulates gene expression and apoptosis. In humans HIPK2 is involved in acute myeloid leukemia and myelodysplastic syndrome [12]. The correlation of HIPK2 expression with GDF11 expression we observed (Figure 3) may indicate HIPK2 modulation of GDF11 effect.

Our analysis above suggests that GDF11 or an analogue might be therapeutic in grade 3 glioma. GDF11 is a tumor suppressor for pancreatic cancer, promoting apoptosis [13], although GDF11 upregulation is associated with reduced survival in uveal melanoma [14]. GDF11 does not cross the blood brain barrier but affects the brain by acting on brain endothelial cells [15]. GDF11 might be delivered to a brain tumor intranasally. Further study would be worthwhile.

## Data Availability

Data publicly available from the Cancer Genome Atlas

https://www.cancer.gov/about-nci/organization/ccg/research/structural-genomics/tcga

